# Supervised Machine-Learning Classification of Treatment-Resistant Depression in U.S. Claims Data

**DOI:** 10.1101/2025.03.26.25324691

**Authors:** Vicki K Wing, Jason Lerner, Patrick Clarke, Shane O’Connor, Sydney Tran, Jonathan Darer, Jaspreet Gill, Lucinda Orsini

## Abstract

**Background:** Accurate identification of individuals with treatment-resistant depression (TRD) is important to facilitate timely access to appropriate care. However, this process currently depends on subjective provider assessments and onerous medication calculations in real-world data. To reduce the burden of TRD identification, we developed TRD classification models using data extracted from a claims database.

**Methods:** Using U.S. commercial claims data, we developed and tested two models using automated machine learning, as well as a rule-based model modified from a published TRD proxy. The highest performing model was designated the full-feature model. The parsimonious model was determined by implementing backward elimination and a clinically oriented consolidation strategy on the full-feature model. The rule-based model was adapted from a published proxy definition.

**Results:** The full feature model (ridge logistic regression) demonstrated the highest overall performance (AUC=0.96, F1=0.64) with 306 features. Backward elimination and implementation of the feature consolidation strategy resulted in a parsimonious model (logistic regression) with acceptable performance (AUC=0.92, F1=0.53) comprising 8 drug-class features. The rule-based model (decision tree) had the lowest AUC (0.82) and F1 score (0.40).

**Conclusions:** To address the need for efficient TRD identification, we developed a parsimonious machine learning model capable of identifying individuals with TRD from claims data based on 8 drug class features. This model performs near the limit established by the full features model and has an interpretable architecture. Furthermore, it can be used to support population health and outcomes research and may reduce the subjectivity and variability in approaches to TRD identification in clinical practice.

## INTRODUCTION

Major depressive disorder (MDD) affects 8.3% of individuals living in the United States (NIMH; Zhdanava et al., 2021). Treatment-resistant depression (TRD), defined by the U.S. Food and Drug Administration and the European Medicines Agency as the failure to respond to two or more antidepressant medication trials of adequate dose and duration, affects an estimated 5.8% to 30% of individuals with MDD (Zhdanava et al., 2021; Liu et al., 2021; Nemeroff, 2007; US FDA). Compared to individuals with MDD who respond to treatment, individuals with TRD have higher rates of hospitalizations with longer inpatients stays, more lost workdays, and higher rates of mortality and self-harm behaviors (Adekkanattu et al., 2023; Lundberg et al., 2023). Estimates of direct and indirect costs of TRD in the United States exceed $43 billion annually (Zhdanava et al., 2021).

Published clinical guidelines for the management of TRD (Bennabi et al., 2019; VA/DoD, 2023) vary with respect to recommended diagnostics, augmentation pharmacotherapy (i.e., lithium, atypical antipsychotics, thyroid hormone, ketamine, esketamine), neuromodulation therapies (i.e., electroconvulsive therapy, repetitive transcranial magnetic stimulation, deep brain stimulation, vagus nerve stimulation), and psychotherapy (Voineskos et al., 2020, Gabriel et al., 2023). However, there is consistent agreement regarding the need for prompt identification of individuals with TRD and escalation of therapy with appropriate monitoring to achieve symptom improvement and resolution (Rybak et al., 2021).

The current dominant clinical definition of TRD incorporates a) a clinician assessment of failure to respond to treatment, b) an understanding of minimum antidepressant therapeutic dosing, and c) an assessment of patient adherence to the antidepressants for a minimum duration to achieve therapeutic effectiveness (Gelenberg et al., 2010). Due to several limitations, this definition is not readily amenable to efficient identification of TRD patients for use in clinical quality improvement initiatives, population-health, and research efforts. First, there is no ICD-10-CM code for TRD, making it difficult to document or analyze the condition in administrative datasets. Second, EHRs may not provide a complete representation of dose and duration for all drugs used to treat depression, given that these may be prescribed and filled outside the EHR system. Third, though operational definitions for TRD have been applied to multiple claims-based datasets, heterogeneity in medication lists, thresholds for adequate dose, and medication duration requirements may lead to disparate conclusions. Finally, since most cases of major depression are managed in primary care settings, assessment of symptom change may be performed by providers with limited experience treating severe depression or without the use of formal assessment instruments (Rizvi et al., 2014). Due to these limitations, the identification of the subset of MDD patients with TRD may be an impediment not only to the advancement of research in TRD, but also to timely referral and treatment for individuals with this condition (Chopra et al., 2023; Simon, 2010). To develop a more practical method of identifying individuals with TRD, we developed a predictive algorithm using supervised machine learning applied to medical and pharmacy insurance claims data.

## METHODS

In this study, we used U.S. commercial insurance claims data to develop and test three machine learning models, including a full feature model, a parsimonious model developed from automated machine learning, and a rule-based definition to classify TRD. The study population was limited to individuals receiving medication for treatment of MDD, and models were trained to distinguish TRD from non-TRD individuals in this population.

### Identification and Selection of Study Participants

Study data were derived from the Merative MarketScan^®^ Commercial Claims and Encounters Database, a longitudinal medical and pharmacy claims dataset for commercially insured individuals across the United States. The total study period was January 1, 2019 to December 31, 2022, with a case identification period beginning December 31, 2020. The index date was defined as the date of an individual’s last pharmacy claim for antidepressant treatment during the case identification period. The observation period, defined as the two-year period prior to the index date, was used to establish participant eligibility, determine TRD or non-TRD groups, and collect feature information.

Eligible participants met each of the following criteria: a) at least one pharmacy claim for an antidepressant during the case identification period, b) at least two medical claims associated with an ICD-10-CM diagnosis code of MDD (F32.X, F33.X, F34.1X) in an outpatient encounter ≤ 1 year apart or one of these diagnoses in an inpatient encounter during the observation period, c) two or more years of continuous pharmacy and medical benefits eligibility preceding the index date, and d) between the ages of 18 and 64 at the start of the observation period. Individuals with one or more medical claims associated with an ICD-10-CM diagnostic code for bipolar disorder, schizophrenia, dementia, or schizoaffective disorder during the observation period, were excluded from the study.

From the dataset, two cohorts were defined for supervised machine learning, a “TRD Cohort” and a “Non-TRD Cohort”. The TRD Cohort included individuals who failed to respond to at least two depression medications of adequate dose for a minimum duration of 6 weeks. Failure was defined as a switch or addition of a depression medication suggestive of ongoing depressive symptoms (Amos et al., 2018). In our study, “adequate dose” for most drugs was sourced from the minimum therapeutic dose for medications listed in the Massachusetts General Hospital Antidepressant Treatment Response Questionnaire (MGH-ATRQ) (Supplemental Table 1). For drugs not referenced in the MGH-ATRQ, “adequate dose” was sourced from product labels. Daily dose was calculated for each medication based on metric quantity, days of medication supplied, and the dose as defined by the national drug code (NDC). The non-TRD Cohort comprised all remaining patients in the study population who did not meet the TRD criteria.

### Modeling and Feature Selection

The study included 501,493 eligible study participants, with 463,485 (92.4%) individuals in the non-TRD Cohort and 38,008 (7.6%) in the TRD Cohort. Individuals were randomly split into cohort balanced 80%/20% training/test sets. The training and test cohorts did not differ significantly in age, insurance type, geographic region, number of unique medication classes, psychiatric comorbidities, or psychiatric visits.

Selection of features to be included in the full feature model was based upon a published systematic review conducted by O’Connor et al. (2023) on risk factors associated with TRD. The review identified several mental health, physical health, demographic, genetic, imaging, personality, and biological risk factors of TRD, including depression severity, suicidality, duration of depressive episodes, and recurrent depression. Common demographic risk factors included younger age and female sex, and common physical health-related risk factors included cardiovascular disease, pain, and thyroid dysfunction (O’Connor et al., 2023).

A total of 517 features were initially identified to be included in the full feature model, which included demographics (e.g., age), psychiatric comorbidities (e.g., social phobia, substance use), physical comorbidities (e.g., pain, diabetes), medication days supplied, drug classes, generic drug names, and mental health and non-mental health resource utilization (e.g., number of emergency department visits or hospital admissions). Using the training set, features were pruned by removing one feature from pairs with correlations greater than 0.9 and those with coefficients of 0 when fit to a LASSO regression. We subsequently trained the automated machine learning (AutoML) engine on the remaining features using H2O.ai version 3.46.0.3 in Python. The AutoML process generated 20 models, including distributed random forest, extremely randomized trees, regularized generalized linear models, Gradient Boosting Machine (GBM), and Extreme Gradient Boosting Machine (XGBoost GBM). Deep learning and stacked ensemble options were excluded due to the significant increase in runtime for these more complex models. Given the strong performance of the ridge logistic regression model, further exploration of more complex models was considered unnecessary. We implemented 5-fold cross-validation within the training data and ranked models according to their F1 scores, with the highest-performing model designated as the “full feature model”.

To develop a more parsimonious model, we examined variable importance scores for the variables in the full feature model, which were calculated within AutoML and indicated relative influence of each variable to the model. Given that the Top 50 most influential variables in the full feature model pertained to MDD medication use, we focused on medication class features which count the number of medications in each class used by a patient during the observation period (i.e., the binned *_UNIQUE_COUNT variables listed in Appendix B).

Using only the 39 medication class features, we implemented a systematic feature selection approach using ridge logistic regression with automatic lambda parameter optimization and 5-fold cross-validation. The feature selection process followed an iterative elimination strategy. The model was trained starting with all drug-class features, and the variable with the lowest variable importance ranking was removed at each step until only one feature remained. At each iteration, model performance was evaluated using the mean F1 score. The F1 score curve for each model was examined to identify the point of diminishing returns, where additional features no longer result in significant improvements in the F1 score. This was determined by observing a marked reduction in the rate of change, indicated by a large finite difference on the curve, also termed the “elbow method”. Specifically, the feature count was determined at the point where the downward slope of the F1 score, as a function of the number of features, began to steepen significantly. The elbow of the curve was interpreted as the threshold where further feature reduction would result in meaningful performance deterioration.

For comparative purposes, we also tested a rule-based definition (decision tree model) based on a modified version of a published TRD proxy (Cepeda et al., 2017). Using medical claims data, Cepeda et al. (2017) used procedure codes for electroconvulsive therapy, deep brain stimulation, or vagus nerve stimulation to indicate a proxy for TRD. and found that use of ≥ 1 antipsychotic and ≥ 1 antidepressant or ≥ 3 antidepressants in the last year effectively discriminated between individuals with and without TRD. Based upon these findings, we developed a decision tree model that predicted TRD based on whether a patient had received either a) three distinct antidepressants or b) one antidepressant and one atypical antipsychotic during the observation period.

## RESULTS

There was no significant difference in age between the TRD and non-TRD cohorts (Table 1). The TRD Cohort had higher rates of moderate to severe MDD compared to the non-TRD Cohort (47.9% vs. 29.7%, SMD = 0.38), as well as higher rates of suicidal ideation (3.2% vs. 1.1%, SMD = 0.14). Rates of psychiatric comorbidities were also higher among the TRD Cohort compared to the non-TRD Cohort, including anxiety (58.9% vs. 45.2%, SMD = 0.28), ADHD (11.4% vs. 8.5%, SMD = 0.10), PTSD (8.8% vs. 4.2%, SMD = 0.19), alcohol abuse and dependence (4.9% vs. 3.0%, SMD = 0.10), and panic disorder (4.8% vs. 2.6%, SMD = 0.12). Overall, the TRD Cohort also had more psychiatric-related healthcare resource utilization over the 2-year observation period than did the non-TRD Cohort in all the following categories: office psychiatric visits (*M* = 3.2 vs. *M* = 1.1, SMD = 0.29), telehealth psychiatric visits (*M* = 1.6 vs. *M* = 0.7, SMD = 0.20), office psychotherapy visits (*M* = 8.7 vs. *M* = 4.8, SMD = 0.25), and telehealth psychotherapy visits (*M* = 5.9 vs. *M* = 3.6, SMD = 0.17) (Supplemental Table 2).

**Table 1.** MDD and TRD Population Characteristics.

|  | TRD<br>(N=38,008)<br>n (%) | MDD<br>(N=463,485)<br>n (%) | SMD |
| --- | --- | --- | --- |
| **Demographics** |  |  |  |
| Age, years |  |  | 0.08 |
| 18-34 | 13222 (34.8) | 155316 (33.5) |  |
| 35-44 | 8860 (23.3) | 100271 (21.6) |  |
| 45-54 | 8984 (23.6) | 109074 (23.5) |  |
| 55-64 | 6942 (18.3) | 98824 (21.3) |  |
| Sex |  |  |  |
| Female |  |  |  |
| **Depression** |  |  |  |
| Recurrent MDD | 13292 (35.0) | 81268 (17.5) | 0.40 |
| Moderate to Severe MDD | 18199 (47.9) | 137664 (29.7) | 0.38 |
| Suicidal Ideation | 1204 (3.2) | 5219 (1.1) | 0.14 |
| Suicidal Attempt | 494 (1.3) | 2161 (0.5) | 0.09 |
| **Other Mental Health Disorders** |  |  |  |
| Anxiety | 22378 (58.9) | 209319 (45.2) | 0.28 |
| ADHD | 4326 (11.4) | 39334 (8.5) | 0.10 |
| PTSD | 3331 (8.8) | 19264 (4.2) | 0.19 |
| Alcohol Abuse Dependence | 1868 (4.9) | 13679 (3.0) | 0.10 |
| Panic Disorder | 1828 (4.8) | 12009 (2.6) | 0.12 |
| Eating Disorders | 885 (2.3) | 5089 (1.1) | 0.10 |
| Drug Abuse | 846 (2.2) | 4384 (0.9) | 0.10 |
| Opioid Dependence | 800 (2.1) | 4959 (1.1) | 0.08 |
| Tobacco Dependence | 638 (1.7) | 6269 (1.4) | 0.03 |
| Social Phobia | 517 (1.4) | 3553 (0.8) | 0.06 |
| Nonorganic Psychosis | 439 (1.2) | 1861 (0.4) | 0.09 |
| Personality Disorder | 104 (0.3) | 471 (0.1) | 0.04 |

### Full-Feature Model

After pruning features using LASSO regression, 306 features remained and were input to the AutoML engine to train and refine the models. A ridge logistic regression model achieved the highest F1 score on the full feature training set. In the test set, the ridge logistic regression model yielded an AUC of 0.96, an F1 score of 0.64, and sensitivity and specificity of 78% and 95%, respectively (Table 2).

**Table 2.** Performance Metrics of the Test Sets.

| Model | AUC | F1 | Sensitivity | Specificity | PPV | NPV | Accuracy |
| --- | --- | --- | --- | --- | --- | --- | --- |
| Full-Feature Model (Ridge Logistic Regression) | 0.96 | 0.64 | 0.78 | 0.95 | 0.54 | 0.95 | 0.93 |
| Parsimonious Model (Logistic Regression) | 0.92 | 0.53 | 0.66 | 0.93 | 0.44 | 0.93 | 0.91 |
| Rule-Based Definition (Decision Tree) | 0.82 | 0.40 | 0.84 | 0.81 | 0.26 | 0.81 | 0.81 |

### Parsimonious Model

An iterative elimination of the 39 medication class features for the parsimonious model indicated an elbow point in performance at 14 features (Figure 2). The features primarily included granular medication usage patterns (e.g., use of two SSRIs, use of one atypical antipsychotics). To enhance clinical applicability and interpretability, these features were consolidated into broader, binary indicators. This transformation turned the features into more practical and clinically actionable categories, while maintaining the essential predictive information in the original model. Consolidation of the variables produced the following 8 features to be included in the parsimonious model: at least 2 SSRIs used (OR 10.23), at least 1 anticonvulsant used (OR 8.81), at least 1 atypical antipsychotic used (OR 7.53), at least 1 NDRI used (OR 7.17), and at least 1 thyroid hormone used (OR 6.95), at least 1 mood stabilizer used (OR 6.14), at least 1 tetracyclic antidepressant used (OR 3.75), and at least 1 SNRI used (OR 3.38) (Table 3).

**Table 3.**
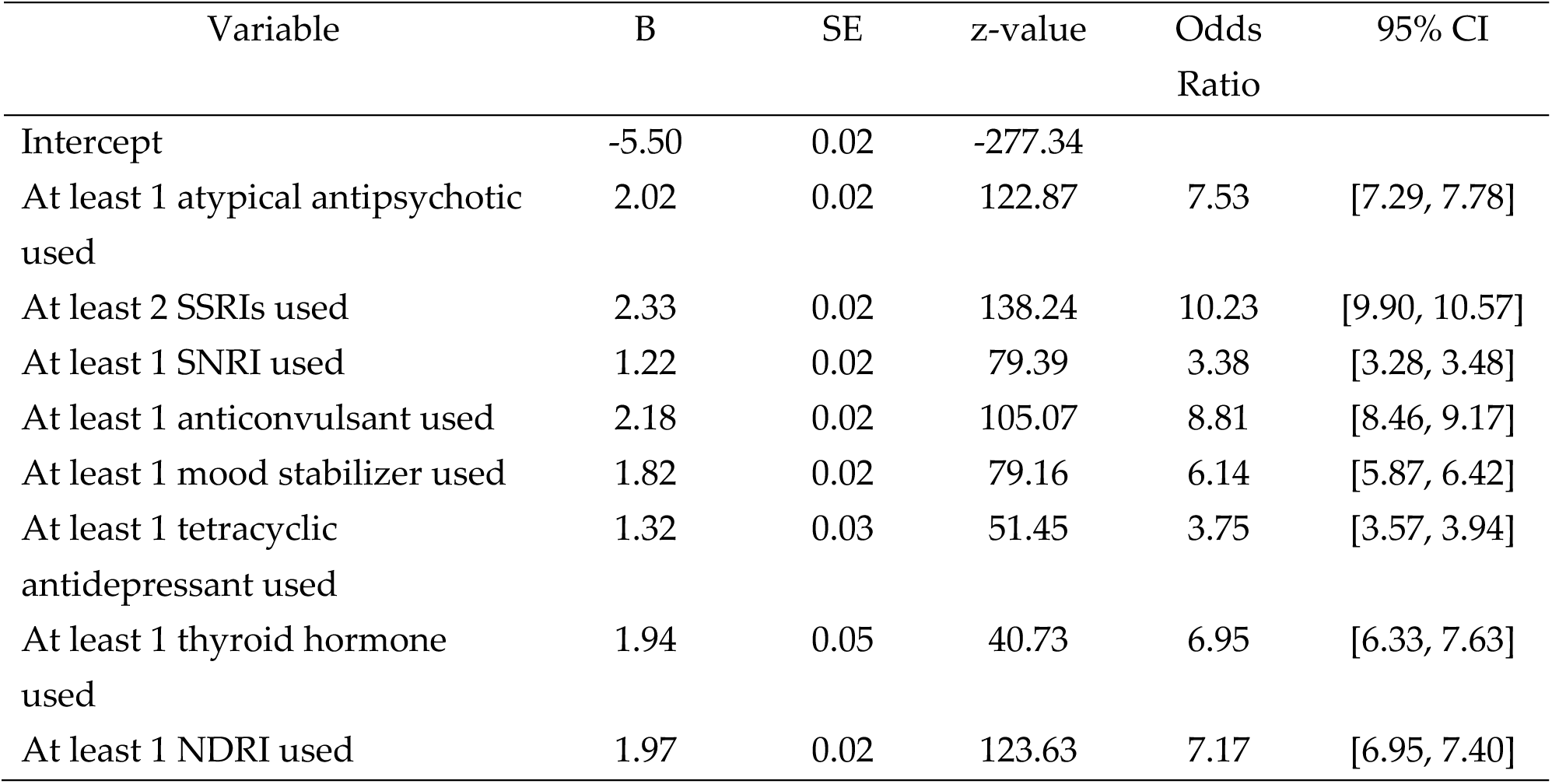
Parsimonious (Logistic Regression) Model Characteristics.

| Variable | B | SE | z-value | Odds Ratio | 95% CI |
| --- | --- | --- | --- | --- | --- |
| Intercept | -5.50 | 0.02 | -277.34 |  |  |
| At least 1 atypical antipsychotic used | 2.02 | 0.02 | 122.87 | 7.53 | [7.29, 7.78] |
| At least 2 SSRIs used | 2.33 | 0.02 | 138.24 | 10.23 | [9.90, 10.57] |
| At least 1 SNRI used | 1.22 | 0.02 | 79.39 | 3.38 | [3.28, 3.48] |
| At least 1 anticonvulsant used | 2.18 | 0.02 | 105.07 | 8.81 | [8.46, 9.17] |
| At least 1 mood stabilizer used | 1.82 | 0.02 | 79.16 | 6.14 | [5.87, 6.42] |
| At least 1 tetracyclic antidepressant used | 1.32 | 0.03 | 51.45 | 3.75 | [3.57, 3.94] |
| At least 1 thyroid hormone used | 1.94 | 0.05 | 40.73 | 6.95 | [6.33, 7.63] |
| At least 1 NDRI used | 1.97 | 0.02 | 123.63 | 7.17 | [6.95, 7.40] |

A standard, non-regularized logistic regression model that was trained using the 8 features was used as the final parsimonious model. Regularization did not significantly change the coefficients, so we used the non-regularized model for simplicity. In the test set, the parsimonious model achieved an AUC of 0.92, an F1 score of 0.53, a sensitivity of 66%, and a specificity of 93% (Figure 1; Table 2).

**Figure 1.**
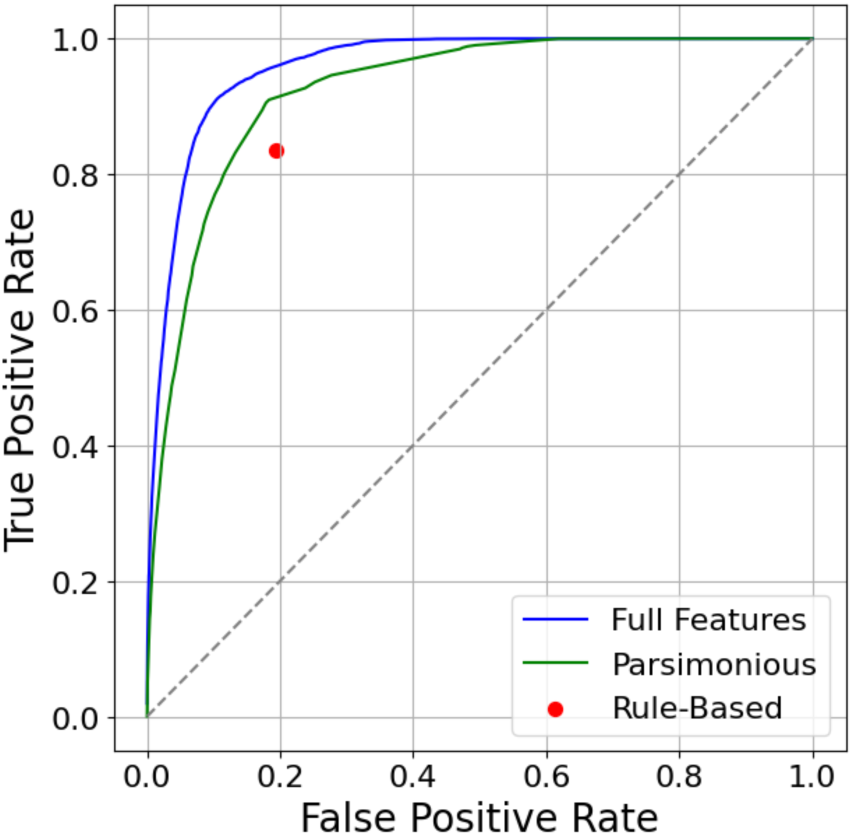
AUCs for the Parsimonious (Logistic Regression), Full Feature, and Rule-Based Models.

**Figure 2.**
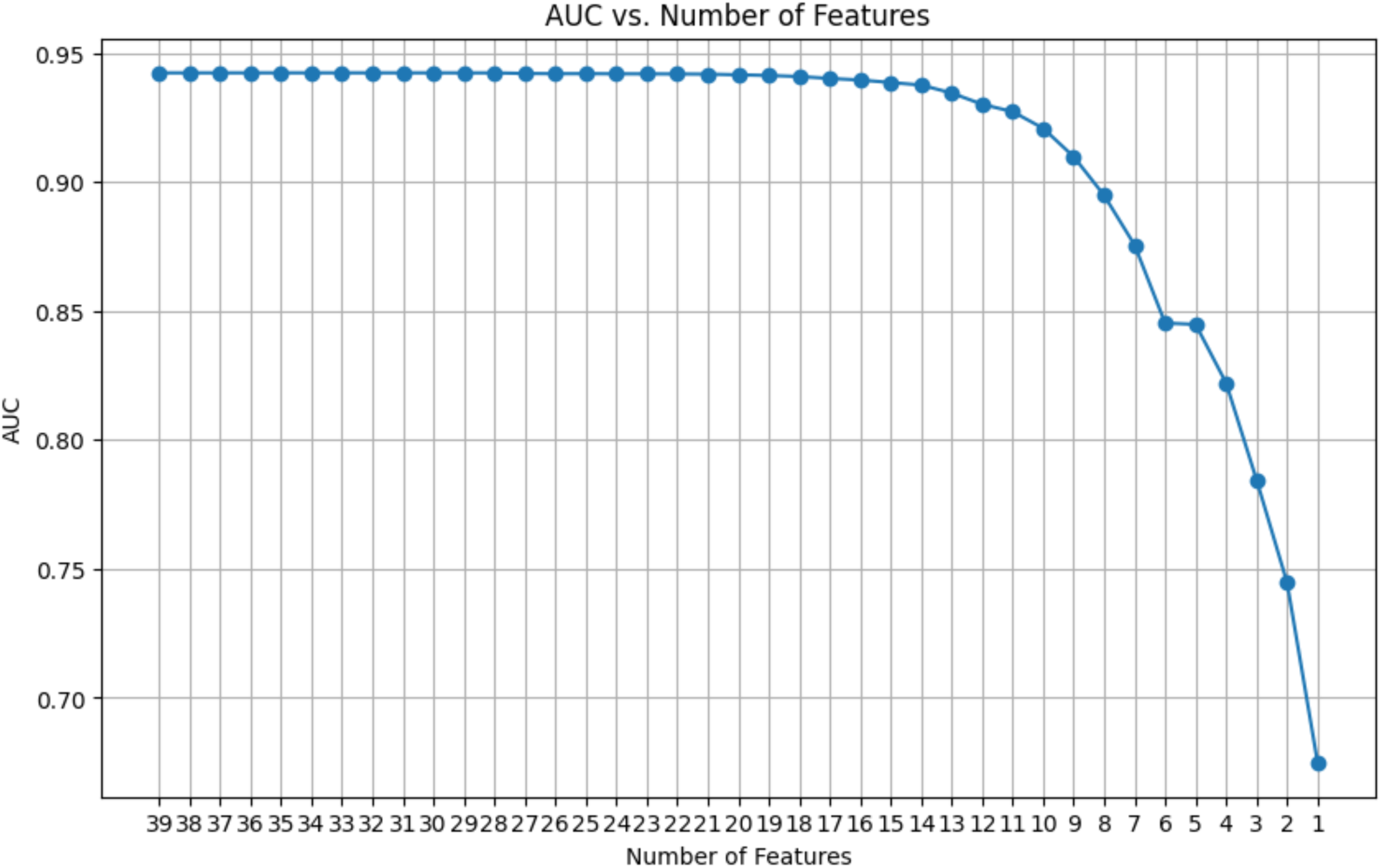
AUC by Descending Number of Features.

### Rule-Based Definition

The decision tree, developed with the proxy TRD feature, yielded acceptable performance metrics, with an AUC of 0.82, an F1 score of 0.40, a sensitivity of 84%, and a specificity of 81% (Table 2).

## DISCUSSION

To reduce the burden and subjectivity of the identification of individuals with TRD for clinical and research purposes, we sought to develop a proxy definition of TRD based upon elements easily available with claims datasets and electronic health records (EHRs). Using supervised machine learning and statistical methods, three models were developed, all with acceptable performance. The parsimonious model comprised 8 data features and demonstrated good utility characteristics without the need for drug dose and duration information.

This pragmatic approach to identifying individuals with TRD in clinical practice is intended to facilitate adherence to best practices, escalate therapy including referral to advanced psychiatric care, and reinforce monitoring. In the absence of a TRD ICD-10-CM diagnosis code, establishing a common set of criteria for the identification of TRD in EHRs and claims data is difficult. EHR vendors embed structured diagnostic vocabularies to support clinical care with mapping to ICD-10-CM codes, but the inclusion of a TRD concept is not well documented. The limitations of TRD-related informatics and reporting highlight the need for simple, efficient methods to identify individuals with TRD at the point-of-care to remind clinicians to consider additional therapies or referrals above and beyond the needs of treatment-sensitive MDD.

The importance of early identification and treatment of individuals with MDD and TRD has implications for monitoring, disease progression, and escalation of care. Individuals with prolonged untreated MDD are more likely to develop TRD, and those with TRD are at increased risk of suicidal ideation and attempted suicide (Orsini et al., 2022; Hun et al, 2017). Individuals with TRD are also at increased risk for comorbid ischemic heart disease, stroke, chronic kidney disease, cancer, as well as increased risk for ED visits and hospitalizations (Adekkanattu et al., 2023). Enabling early and efficient identification of individuals with TRD can facilitate escalation in treatment.

Application of machine learning methods to TRD is an evolving field. Published machine learning models have sought to predict risk for TRD as well as potential response to therapy (Kautzky et al, 2017; Perlis, 2013; Lage et al., 2022; Pigoni et al., 2019). These advanced TRD-related risk prediction models show promise, but their utility is, in part, dependent upon a health system’s ability to efficiently and effectively identify individuals with TRD within their EHRs.

## STRENGTHS

This study made use of administrative claims data including total daily dose of antidepressant treatment to establish a simplified 8-element model for the identification of individuals with TRD. The use of administrative insurance claims data enables this model to be efficiently translated to other claims datasets. Additionally, the model includes 8 elements which are all easily available through EHR and/or claims-based data sources, enabling application within clinical and population-health settings.

For purposes of research, the above models can enable efficient identification of individuals with TRD in nationally representative administrative claims datasets to assess practice patterns and healthcare utilization. The parsimonious model not only requires minimal data inputs but also demonstrated excellent overall classification performance (AUC and F1). Further, the logistic regression model has inherent advantages for application within distinct use cases (e.g., escalation of therapy, referral to TRD expert care, and research). In particular, the model’s classification threshold for the probability of TRD can be varied depending on the cost of inaccurate classification (e.g., tradeoffs between type I and type II errors) within each use case.

## LIMITATIONS

This study is subject to the limitations of all administrative claims-based research with incomplete coding and potential for misclassification of populations. Additionally, while the use of two trials of antidepressants and a third agent suggested initial failure and the persistence of depressive symptoms, there is no clinical confirmation that individuals failed to respond. The use of commercial administrative claims dataset used to develop the models introduces potential bias and limits the generalizability to all patient populations (e.g., Medicaid, Medicare). Furthermore, the use of data from 2019-2022 during the COVID-19 pandemic may also limit generalizability to non-pandemic periods.

As with any predictive model, the models developed in this study require robust validation within a clinical context to ascertain veracity and generalizability across disparate populations. These efforts will be critical to assess ease of use by researchers, at the point of care, and for population health initiatives.

## CONCLUSIONS

We developed a machine learning classifier to identify individuals with TRD using a simplified set of data elements existing within EHRs and insurance claims data. The classifier was characterized by high performance, supporting its use in research which may facilitate the advance of care of individuals with TRD. It also serves as an important first step towards a clinically useful model.

## Data Availability

All data produced in the present study are available upon reasonable request to the authors

## Acknowledgements

The authors would like to thank ML Stinstrom for her administrative support.

---

**Supplemental Table 1.**
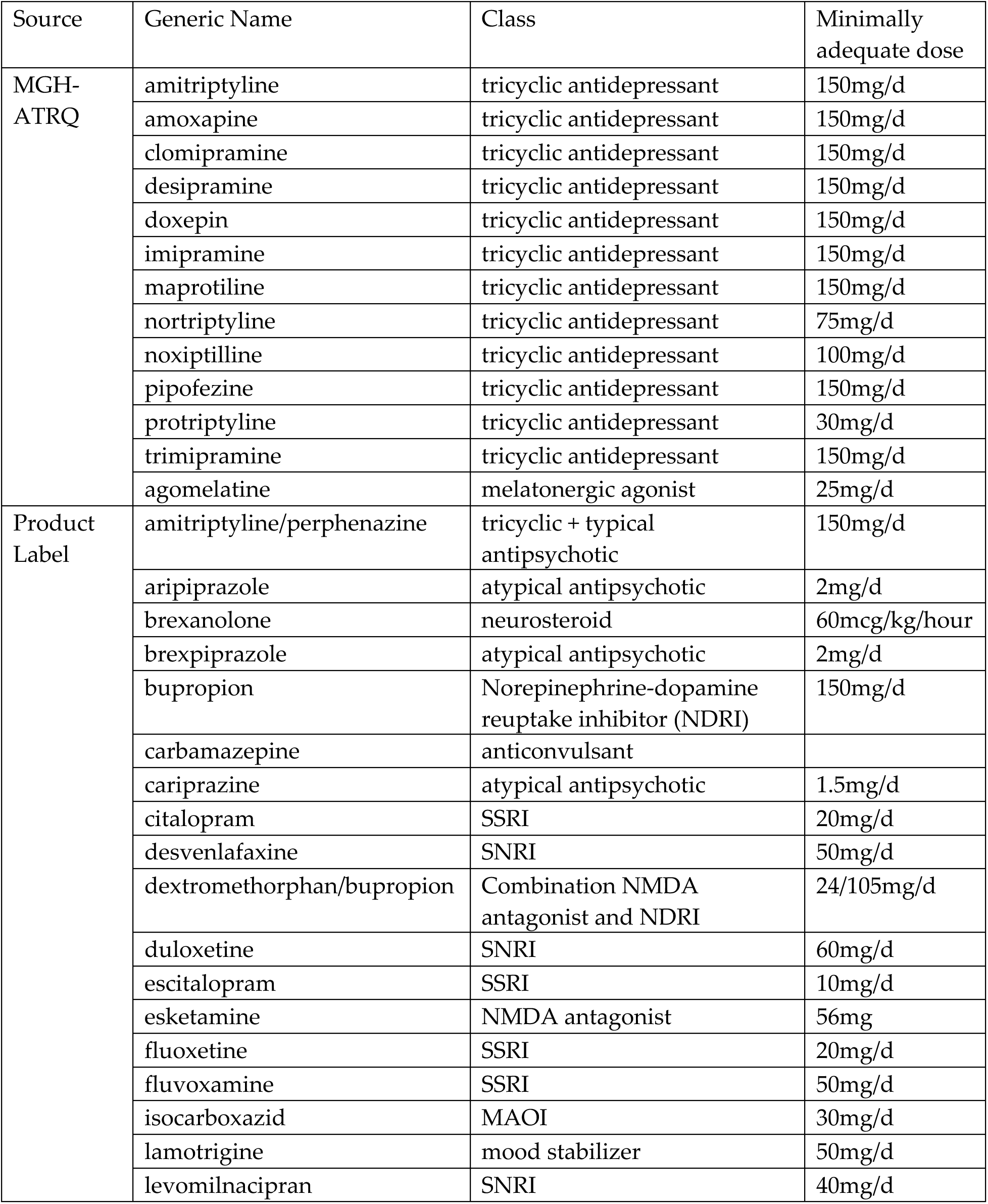
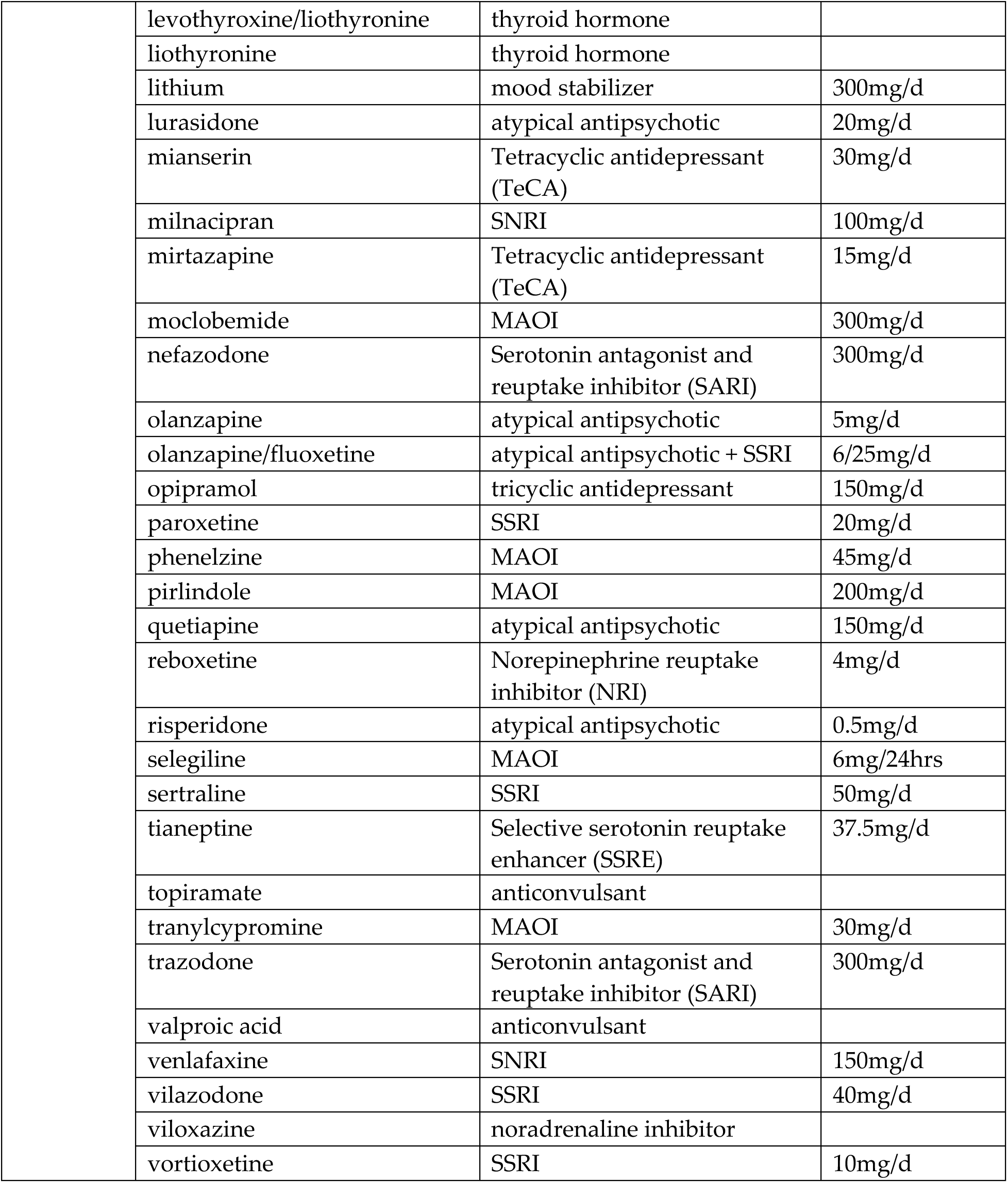
Minimally Adequate Dose for Medication Classes.

**Supplemental Table 2.** Demographic Comparisons Between Training and Test Cohorts.

| Domain | Variable | Overall | Training | Test | SMD<sup>a</sup> |
| --- | --- | --- | --- | --- | --- |
| Age<sup>b</sup> | Age | 41.3 (13.4) | 41.3 (13.4) | 41.3 (13.3) | -0.00 |
| Insurance Plan Type<sup>c</sup> | PPO | 279,625 (55.8) | 223,976 (55.8) | 55,649 (55.6) | 0.01 |
|  | CDHP/HDHP | 120,039 (23.9) | 96,032 (23.9) | 24,007 (24) |  |
|  | HMO | 76,917 (15.3) | 61,390 (15.3) | 15,527 (15.5) |  |
|  | Comprehensive | 16,132 (3.2) | 12,971 (3.2) | 3,163 (3.2) |  |
|  | None | 5,385 (1.1) | 4,302 (1.1) | 1083 (1.1) |  |
|  | EPO | 3,384 (0.7) | 2,694 (0.7) | 690 (0.7) |  |
|  | Basic/Major Medical | 9 (0) | 5 (0) | 4 (0) |  |
| Geographic Region<sup>c</sup> | North Central | 131,789 (26.3) | 105,385 (26.3) | 26,404 (26.4) | 0.01 |
|  | Northeast | 58,503 (11.7) | 46,816 (11.7) | 11,687 (11.7) |  |
|  | South | 237,285 (47.3) | 190,133 (47.4) | 47,152 (47.1) |  |
|  | Unknown | 1,076 (0.2) | 880 (0.2) | 196 (0.2) |  |
|  | West | 72,840 (14.5) | 58,156 (14.5) | 14,684 (14.7) |  |
| Number of Unique Medications<sup>b</sup> | Other Antidepressants | 0.5 (0.6) | 0.5 (0.6) | 0.5 (0.6) | 0.00 |
|  | SSRIs | 0.9 (0.7) | 0.9 (0.7) | 0.9 (0.7) | 0.00 |
|  | SNRIs | 0.3 (0.5) | 0.3 (0.5) | 0.3 (0.5) | 0.00 |
|  | Mood Stabilizers | 0.0 (0.2) | 0.0 (0.2) | 0.0 (0.2) | 0.00 |
|  | Atypical Antipsychotics | 0.1 (0.4) | 0.1 (0.4) | 0.1 (0.4) | -0.00 |
|  | Tricyclics | 0.1 (0.3) | 0.1 (0.3) | 0.1 (0.3) | 0.00 |
|  | Anticonvulsants | 0.1 (0.2) | 0.1 (0.2) | 0.1 (0.2) | -0.00 |
|  | Thyroid Hormones | 0.0 (0.1) | 0.0 (0.1) | 0.0 (0.1) | -0.00 |
|  | Atypical Antipsychotic + SSRIs | 0.0 (0.0) | 0.0 (0.0) | 0.0 (0.0) | 0.00 |
|  | Noradrenaline Inhibitors | 0.0 (0.0) | 0.0 (0.0) | 0.0 (0.0) | -0.00 |
|  | NMDA Antagonists | 0.0 (0.0) | 0.0 (0.0) | 0.0 (0.0) | 0.00 |
|  | MAOIs | 0.0 (0.0) | 0.0 (0.0) | 0.0 (0.0) | 0.00 |
|  | Tricyclic + Typical Antipsychotics | 0.0 (0.0) | 0.0 (0.0) | 0.0 (0.0) | 0.00 |
|  | Neurosteroids | 0.0 (0.0) | 0.0 (0.0) | 0.0 (0.0) | 0.00 |
|  | NMDA NDRI | 0.0 (0.0) | 0.0 (0.0) | 0.0 (0.0) | 0.00 |
|  | NDRIs | 0.3 (0.5) | 0.3 (0.5) | 0.3 (0.5) | 0.00 |
|  | SARIs | 0.2 (0.4) | 0.2 (0.4) | 0.2 (0.4) | -0.00 |
|  | TECAs | 0.0 (0.2) | 0.0 (0.2) | 0.0 (0.2) | 0.00 |
| Psychiatric Healthcare Resource Utilization<sup>b</sup> | Inpatient Psychiatric Visits | 0.1 (1.7) | 0.1 (1.6) | 0.2 (1.9) | -0.00 |
|  | Office Psychiatric Visits | 1.2 (5.4) | 1.2 (5.4) | 1.2 (5.4) | 0.00 |
|  | Telehealth Psychiatric Visits | 0.8 (3.7) | 0.8 (3.7) | 0.8 (3.6) | 0.00 |
|  | Hospital Outpatient Psychiatric Visits | 0.1 (1.2) | 0.1 (1.2) | 0.1 (1.3) | -0.00 |
|  | Emergency Department Psychiatric Visits | 0.1 (0.5) | 0.1 (0.5) | 0.1 (0.5) | 0.00 |
|  | Office Psychotherapy Visits | 5.0 (13.5) | 5.1 (13.5) | 5.0 (13.6) | 0.00 |
|  | Telehealth Psychotherapy Visits | 3.7 (11.8) | 3.8 (11.8) | 3.7 (11.7) | 0.00 |

<sup>a</sup>Test, Training. <sup>b</sup>Mean (SD). <sup>c</sup>n (%).

**Supplemental Table 3.** Variable Importance Scores for the 14-Feature Model.

| Variable | Relative Importance | Scaled Importance | Percentage |
| --- | --- | --- | --- |
| 2 SSRIs supplied over 2 years | 1.036 | 1.000 | 0.149 |
| 1 NDRI supplied over 2 years | 1.003 | 0.968 | 0.144 |
| 1 SSRI supplied over 2 years | 0.783 | 0.756 | 0.113 |
| 1 SNRI supplied over 2 years | 0.623 | 0.602 | 0.090 |
| 1 atypical antipsychotic supplied over 2 years | 0.544 | 0.546 | 0.078 |
| 3 SSRIs supplied over 2 years | 0.525 | 0.507 | 0.075 |
| 1 anticonvulsant supplied over 2 years | 0.523 | 0.505 | 0.075 |
| 1 mood stabilizer supplied over 2 years | 0.371 | 0.358 | 0.053 |
| 1 atypical antipsychotic supplied over 2 years | 0.353 | 0.341 | 0.051 |
| 2 SNRI supplied over 2 years | 0.337 | 0.326 | 0.048 |
| 1 TECA supplied over 2 years | 0.277 | 0.268 | 0.040 |
| 1 thyroid hormone supplied over 2 years | 0.202 | 0.195 | 0.030 |
| 4 SSRIs supplied over 2 years | 0.196 | 0.189 | 0.028 |
| 3 atypical antipsychotics supplied over 2 years | 0.181 | 0.175 | 0.026 |

## REFERENCES

Al-harbi, K,S. (2012). Treatment-resistant depression: Therapeutic trends, challenges, and Future Directions. Patient Preference and Adherence, 369. 10.2147/ppa.s29716

Adekkanattu P, Olfson M, Susser LC, Patra B, Vekaria V, Coombes BJ, Lepow L, Fennessy B, Charney A, Ryu E, Miller KD, Pan L, Yangchen T, Talati A, Wickramaratne P, Weissman M, Mann J, Biernacka JM, Pathak J. Comorbidity and healthcare utilization in patients with treatment resistant depression: A large-scale retrospective cohort analysis using electronic health records. J Affect Disord. 2023 Mar 1;324:102–113. doi: 10.1016/j.jad.2022.12.044. Epub 2022 Dec 15. PMID: 36529406; PMCID: PMC10327872.

Amos T.B., Tandon, N., Lefebvre, P., Pilon, D., Kamstra, R. L., Pivneva, I., & Greenberg, P.E. (2018). Direct and indirect cost burden and change of employment status in Treatment-Resistant Depression: A matched-cohort study using a US commercial claims database. Journal of Clinical Psychiatry. 79(2):5360. doi: https://10.4088/JCP.17m11725

Andrilla, C. H., Patterson, D. G., Garberson, L. A., Coulthard, C., & Larson, E. H. (2018). Geographic variation in the supply of selected Behavioral Health Providers. American Journal of Preventive Medicine, 54(6). 10.1016/j.amepre.2018.01.004

Baudot, J., Soeiro, T., Tambon, M., Navarro, N., Veyrac, G., Mezaache, S., & Micallef, J. (2021). Safety concerns on the abuse potential of Esketamine: Multidimensional Analysis of a new anti-depressive drug on the market. Fundamental & Clinical Pharmacology, 36(3), 572–581. 10.1111/fcp.12745

Bennabi D, Charpeaud T, Yrondi A, Genty JB, Destouches S, Lancrenon S, Alaïli N, Bellivier F, Bougerol T, Camus V, Dorey JM, Doumy O, Haesebaert F, Holtzmann J, Lançon C, Lefebvre M, Moliere F, Nieto I, Rabu C, Richieri R, Schmitt L, Stephan F, Vaiva G, Walter M, Leboyer M, El-Hage W, Llorca PM, Courtet P, Aouizerate B, Haffen E. Clinical guidelines for the management of treatment-resistant depression: French recommendations from experts, the French Association for Biological Psychiatry and Neuropsychopharmacology and the fondation FondaMental. BMC Psychiatry. 2019 Aug 28;19(1):262. doi: 10.1186/s12888-019-2237-x. PMID: 31455302; PMCID: PMC6712810. BMJ https://ebm.bmj.com/content/18/1/5

Brenes, G. A., Danhauer, S. C., Lyles, M. F., Hogan, P. E., & Miller, M. E. (2015). Barriers to mental health treatment in rural older adults. The American Journal of Geriatric Psychiatry, 23(11), 1172–1178. 10.1016/j.jagp.2015.06.002

Cepeda M.S., Reps, J., Fife, D., Blacketer, C., Stang, P., Ryan, P. (2018) Finding treatment-resistant depression in real-world data: How a data-driven approach compares with expert-based heuristics. Depression and Anxiety, 35(3), 220–228. doi: https://10.1002/da.22705

Center for Disease Control Public Health Information Network Vocabulary Access and Distribution System (PHIN VADS) (2024). https://phinvads.cdc.gov/vads/ViewCodeSystemConcept.action?oid=2.16.840.1.113883.6. 96&code=35489007

Charlson, M. E., Pompei, P., Ales, K. L., & MacKenzie, C. R. (1987). A new method of classifying prognostic comorbidity in Longitudinal Studies: Development and validation. Journal of Chronic Diseases, 40(5), 373–383. 10.1016/0021-9681(87)90171-8

Chopra A, Luccarelli J, Cohen JN, Mischoulon D, Stern TA. Evaluation, Treatment, and Referral of Treatment-Resistant Depression in Primary Care. Prim Care Companion CNS Disord. 2023 Jul 25;25(4):22f03438. doi: 10.4088/PCC.22f03438. PMID: 37506395; PMCID: PMC10564558.

Davis, A. K., Agin-Liebes, G., España, M., Pilecki, B., & Luoma, J. (2021). Attitudes and beliefs about the therapeutic use of psychedelic drugs among psychologists in the United States. Journal of Psychoactive Drugs, 54(4), 309–318. 10.1080/02791072.2021.1971343

Drapalski, A. L., Milford, J., Goldberg, R. W., Brown, C. H., & Dixon, L. B. (2008). Perceived barriers to medical care and mental health care among veterans with serious mental illness. Psychiatric Services, 59(8), 921–924. 10.1176/ps.2008.59.8.921

Eberhardt, M. S., & Pamuk, E. R. (2004). The importance of place of residence: Examining health in rural and nonrural areas. American Journal of Public Health, 94(10), 1682–1686. 10.2105/ajph.94.10.1682

Elnitsky, C. A., Andresen, E. M., Clark, M. E., McGarity, S., Hall, C. G., & Kerns, R. D. (2013). Access to the US Department of Veterans Affairs Health System: Self-reported barriers to care among returnees of Operations Enduring Freedom and iraqi freedom. BMC Health Services Research, 13(1). 10.1186/1472-6963-13-498

Gabriel FC, Stein AT, Melo DO, Fontes-Mota GCH, Dos Santos IB, Rodrigues CDS, Dourado A, Rodrigues MC, Fráguas R, Florez ID, Correia DT, Ribeiro E. Guidelines’ recommendations for the treatment-resistant depression: A systematic review of their quality. PLoS One. 2023 Feb 6;18(2):e0281501. doi: 10.1371/journal.pone.0281501. PMID: 36745622; PMCID: PMC9901785.

Gelenberg, A. J., Freeman, M. P., Markowitz, J. C., Rosenbaum, J. F., Thase, M. E., Trivedi, M. H., & Van Rhoads, R. S. (2010). American Psychiatric Association practice guidelines for the treatment of patients with major depressive disorder. Am J Psychiatry, 167(Suppl 10), 9–118.

Goodwin, G. M., Aaronson, S. T., Alvarez, O., Arden, P. C., Baker, A., Bennett, J. C., Bird, C., Blom, R. E., Brennan, C., Brusch, D., Burke, L., Campbell-Coker, K., Carhart-Harris, R., Cattell, J., Daniel, A., DeBattista, C., Dunlop, B. W., Eisen, K., Feifel, D., … Malievskaia, E. (2022). Single-dose psilocybin for a treatment-resistant episode of Major Depression. New England Journal of Medicine, 387(18), 1637–1648. 10.1056/nejmoa2206443

Hoyt, D. R., Conger, R. D., Valde, J. G., & Weihs, K. (1997). Psychological distress and help seeking in rural America. American Journal of Community Psychology, 25(4), 449–470. 10.1023/a:1024655521619

Hung CI, Liu CY, Yang CH. Untreated duration predicted the severity of depression at the two-year follow-up point. PLoS One. 2017 Sep 21;12(9):e0185119. doi: 10.1371/journal.pone.0185119. PMID: 28934289; PMCID: PMC5608308.

Ionescu, D. F., Rosenbaum, J. F., & Alpert, J. E. (2015). Pharmacological approaches to the challenge of treatment-resistant depression. Dialogues in Clinical Neuroscience, 17(2), 111–126. 10.31887/dcns.2015.17.2/dionescu

Lage I, McCoy TH Jr, Perlis RH, Doshi-Velez F. Efficiently identifying individuals at high risk for treatment resistance in major depressive disorder using electronic health records. J Affect Disord. 2022 Jun 1;306:254–259. doi: 10.1016/j.jad.2022.02.046. Epub 2022 Feb 16. PMID: 35181388; PMCID: PMC9980713.

Leong F.T., Kalibatseva Z.(2011) Cross-cultural barriers to mental health services in the United States. Cerebrum. PMID: 23447774; PMCID: PMC3574791.

Liberman, J., Pesa, J., Rui, P., Joshi, K., & Harding, L. (2023). Social determinants and distance from certified treatment centers are associated with initiation of esketamine nasal spray among patients with challenging-to-treat major depressive disorder. Medicine, 102(7). 10.1097/md.0000000000032895

Liu, X., Mukai, Y., Furtek, C. I., Bortnichak, E. A., Liaw, K.-L., & Zhong, W. (2021). Epidemiology of treatment-resistant depression in the United States. The Journal of Clinical Psychiatry, 83(1). 10.4088/jcp.21m13964

Lundberg, J., Cars, T., Lööv, S.-Å., Söderling, J., Sundström, J., Tiihonen, J., Leval, A., Gannedahl, A., Björkholm, C., Själin, M., & Hellner, C. (2023). Association of treatment-resistant depression with patient outcomes and health care resource utilization in a population-wide study. JAMA Psychiatry, 80(2), 167. 10.1001/jamapsychiatry.2022.3860

McGuire, T. G., & Miranda, J. (2008). New evidence regarding racial and ethnic disparities in Mental Health: Policy Implications. Health Affairs, 27(2), 393–403. 10.1377/hlthaff.27.2.393

McIntyre, R. S., Alsuwaidan, M., Baune, B. T., Berk, M., Demyttenaere, K., Goldberg, J. F., Gorwood, P., Ho, R., Kasper, S., Kennedy, S. H., Ly-Uson, J., Mansur, R. B., McAllister-Williams, R. H., Murrough, J. W., Nemeroff, C. B., Nierenberg, A. A., Rosenblat, J. D., Sanacora, G., Schatzberg, A. F., … Maj, M. (2023a). Treatment-resistant depression: Definition, prevalence, detection, management, and investigational interventions. World Psychiatry, 22(3), 394–412. 10.1002/wps.21120

McIntyre, R. S., Rosenblat, J. D., Nemeroff, C. B., Sanacora, G., Murrough, J. W., Berk, M., Brietzke, E., Dodd, S., Gorwood, P., Ho, R., Iosifescu, D. V., Lopez Jaramillo, C., Kasper, S., Kratiuk, K., Lee, J. G., Lee, Y., Lui, L. M. W., Mansur, R. B., Papakostas, G. I., … Stahl, S. (2021). Synthesizing the evidence for ketamine and esketamine in treatment-resistant depression: An international expert opinion on the available evidence and implementation. American Journal of Psychiatry, 178(5), 383–399. 10.1176/appi.ajp.2020.20081251

Mojtabai, R., Olfson, M., Sampson, N. A., Jin, R., Druss, B., Wang, P. S., Wells, K. B., Pincus, H. A., & Kessler, R. C. (2010). Barriers to mental health treatment: Results from the National Comorbidity Survey Replication. Psychological Medicine, 41(8), 1751–1761. 10.1017/s0033291710002291

Morales, D. A., Barksdale, C. L., & Beckel-Mitchener, A. C. (2020). A call to action to address rural mental health disparities. Journal of Clinical and Translational Science, 4(5), 463–467. 10.1017/cts.2020.42

National Alliance on Mental Illness. (n.d.-a). Continuing disparities in access to mental and physical … https://www.nami.org/Support-Education/Publications-Reports/Public-Policy-Reports/The-Doctor-is-Out/DoctorIsOut

Nemeroff CB. Prevalence and management of treatment-resistant depression. (2007) Journal of Clinical Psychiatry; 68 Suppl 8:17–25. PMID: 17640154.

NIMH: https://www.nimh.nih.gov/health/statistics/major-depression

Noh, J.-W., Kwon, Y. D., Park, J., Oh, I.-H., & Kim, J. (2016). Relationship between physical disability and depression by gender: A panel regression model. PLOS ONE, 11(11). 10.1371/journal.pone.0166238

O’Connor, S. J., Hewitt, N., Kuc, J., & Orsini, L. (2023). Predictors and risk factors of treatment-resistant depression: A systematic review. Journal of Clinical Psychiatry, 85(1), 23r14885.

Orsini LS, O’Connor SJ, Mohwinckel MT, Marwood L, Pahwa AS, Bryder MN, Dong X, Levine SP. Observational study to characterize treatment-resistant depression in Germany, France and the United Kingdom: analysis of real-world data collected through a survey of healthcare professionals. Curr Med Res Opin. 2022 Dec;38(12):2219–2226. doi: 10.1080/03007995.2022.2113692. Epub 2022 Sep 20. PMID: 36106382.

Office of the Commissioner: FDA approves new nasal spray medication for treatment-resistant depression; available only at a certified doctor’s office or clinic. US Food and Drug Administration, 2019 https://www.fda.gov/news-events/press-announcements/fdaapproves-new-nasal-spray-medication-treatment-resistant-depressionavailable-only-certified

Overview. USDA ERS - Rural Poverty & Well-Being. (n.d.). https://www.ers.usda.gov/topics/rural-economy-population/rural-poverty-well-being/

Packness, A., Waldorff, F. B.,Christensen, R. dePont, Hastrup, L. H., Simonsen, E., Vestergaard, M., & Halling, A. (2017). Impact of socioeconomic position and distance on Mental Health Care Utilization: A nationwide Danish follow-up study. Social Psychiatry and Psychiatric Epidemiology, 52(11), 1405–1413. 10.1007/s00127-017-1437-2

Perlis RH. A clinical risk stratification tool for predicting treatment resistance in major depressive disorder. Biol Psychiatry. 2013 Jul 1;74(1):7–14. doi: 10.1016/j.biopsych.2012.12.007. Epub 2013 Feb 4. PMID: 23380715; PMCID: PMC3690142.

Pigoni A, Delvecchio G, Madonna D, Bressi C, Soares J, Brambilla P. Can Machine Learning help us in dealing with treatment resistant depression? A review. J Affect Disord. 2019 Dec 1;259:21–26. doi: 10.1016/j.jad.2019.08.009. Epub 2019 Aug 13. PMID: 31437696.

Probst J,C., Laditka S,B., Moore C,G., Harun N., Powell M,P., Baxley E,G. (2006) Rural-urban differences in depression prevalence: implications for family medicine. Family Medicine. 38(9):653–60. PMID: 17009190.

Raison, C. L., Sanacora, G., Woolley, J., Heinzerling, K., Dunlop, B. W., Brown, R. T., Kakar, R., Hassman, M., Trivedi, R. P., Robison, R., Gukasyan, N., Nayak, S. M., Hu, X., O’Donnell, K. C., Kelmendi, B., Sloshower, J., Penn, A. D., Bradley, E., Kelly, D. F., … Griffiths, R. R. (2023). Single-dose psilocybin treatment for major depressive disorder. JAMA, 330(9), 843. 10.1001/jama.2023.14530

Rizvi SJ, Grima E, Tan M, Rotzinger S, Lin P, McIntyre RS, Kennedy SH. Treatment-Resistant Depression in Primary Care Across Canada. Can J Psychiatry. 2014 Jul;59(7):349–357. doi: 10.1177/070674371405900702. PMID: 25007419; PMCID: PMC4086317

Rybak YE, Lai KSP, Ramasubbu R, Vila-Rodriguez F, Blumberger DM, Chan P, Delva N, Giacobbe P, Gosselin C, Kennedy SH, Iskandar H, McInerney S, Ravitz P, Sharma V, Zaretsky A, Burhan AM. Treatment-resistant major depressive disorder: Canadian expert consensus on definition and assessment. Depress Anxiety. 2021 Apr;38(4):456–467. doi: 10.1002/da.23135. Epub 2021 Feb 2. PMID: 33528865; PMCID: PMC8049072.

Sareen, J., Jagdeo, A., Cox, B. J., Clara, I., ten Have, M., Belik, S.-L., de Graaf, R., & Stein, M. B. (2007). Perceived barriers to mental health service utilization in the United States, Ontario, and the Netherlands. Psychiatric Services, 58(3), 357–364. 10.1176/ps.2007.58.3.357

Serafini, G., Howland, R., Rovedi, F., Girardi, P., & Amore, M. (2014). The role of ketamine in treatment-resistant depression: A systematic review. Current Neuropharmacology, 12(5), 444–461. 10.2174/1570159x12666140619204251

Simmons, L. A., Braun, B., Charnigo, R., Havens, J. R., & Wright, D. W. (2008). Depression and poverty among rural women: A relationship of social causation or social selection? The Journal of Rural Health, 24(3), 292–298. 10.1111/j.1748-0361.2008.00171.x

Simon GE, Perlis RH. Personalized medicine for depression: can we match patients with treatments? Am J Psychiatry. 2010 Dec;167(12):1445–55. doi: 10.1176/appi.ajp.2010.09111680. Epub 2010 Sep 15. PMID: 20843873; PMCID: PMC3723328.

Substance Abuse and Mental Health Services. (n.d.). Racial/Ethnic Differences in Mental Health Service Use among Adults. https://www.samhsa.gov/data/sites/default/files/MHServicesUseAmongAdults/MHServ icesUseAmongAdults.pdf

Swainson, J., Thomas, R. K., Archer, S., Chrenek, C., MacKay, M.-A., Baker, G., Dursun, S., Klassen, L. J., Chokka, P., & Demas, M. L. (2019). Esketamine for Treatment Resistant Depression. Expert Review of Neurotherapeutics, 19(10), 899–911. 10.1080/14737175.2019.1640604

Syed, S. T., Gerber, B. S., & Sharp, L. K. (2013). Traveling towards disease: Transportation Barriers to Health Care Access. Journal of Community Health, 38(5), 976–993. 10.1007/s10900-013-9681-1

US Census Bureau https://www.census.gov/library/stories/2017/08/rural-america.html

US FDA U.S. Food and Drug Administration. Center for Drug Evaluation and Research. Major depressive disorder: developing drugs for treatment. Silver Spring: U.S. Food and Drug Administration, 2018.).

VA/DoD Clinical Practice Guideline. (2022). The Management of Major Depressive Disorder. Washington, DC: U.S. Government Printing Office.

Voineskos, D., Daskalakis, Z. J., & Blumberger, D. M. (2020). Management of treatment-resistant depression: Challenges and strategies. Neuropsychiatric Disease and Treatment, Volume 16, 221–234. 10.2147/ndt.s198774

Walker, E. R., Cummings, J. R., Hockenberry, J. M., & Druss, B. G. (2015). Insurance status, use of mental health services, and unmet need for mental health care in the United States. Psychiatric Services, 66(6), 578–584. 10.1176/appi.ps.201400248

Zhao, G., Okoro, C. A., Hsia, J., Garvin, W. S., & Town, M. (2019). Prevalence of disability and disability types by urban–rural county classification—U.S., 2016. American Journal of Preventive Medicine, 57(6), 749–756. 10.1016/j.amepre.2019.07.022

Zhdanava, M., Kuvadia, H., Joshi, K., Daly, E., Pilon, D., Rossi, C., Morrison, L., Lefebvre, P., & Nelson, C. (2020). Economic burden of treatment-resistant depression in privately insured U.S. patients with physical conditions. Journal of Managed Care; Specialty Pharmacy, 26(8), 996–1007. 10.18553/jmcp.2020.20017

Zhdanava, M., Pilon, D., Ghelerter, I., Chow, W., Joshi, K., Lefebvre, P., & Sheehan, J. J. (2021). The prevalence and national burden of treatment-resistant depression and major depressive disorder in the United States. The Journal of Clinical Psychiatry, 82(2). 10.4088/jcp.20m13699

